# Altered plasma metabolites as fingerprint of cardiac bypass surgery

**DOI:** 10.1101/2025.04.11.25325708

**Authors:** Frieder Neu, Max Wacker, Sven Schuchardt, Sam Vargese, George Awad, Jens Wippermann, Frank Peßler, Priya Veluswamy

## Abstract

**Aims:** Cardiac surgery leads to major post-operative changes in metabolism, but their exact nature and the underlying risk factors remains obscure. We aimed to characterize changes in plasma metabolites after coronary artery bypass grafting (CABG) to identify intra- and post-operative risk factors for global and specific alterations in plasma metabolites post-operatively.

**Methods:** We performed a targeted metabolomic screen on plasma samples from patients undergoing on-pump CABG for coronary artery disease (CAD) (n=24), collected 1 day before surgery and on post-op days 1, 3, and 7. We assessed correlations with parameters of intra-operative course (cardiopulmonary bypass time and aortic cross-clamping time), intensive care unit (ICU) care, (length of ICU stay, duration of mechanical ventilation, duration of epinephrine/dobutamine or norepinephrine therapy), and systemic inflammation.

**Results:** Of the potentially detectable 1019 analytes, 970 passed the quality screen and were included in the analysis. With respect to d0, the greatest degree of change in metabolite populations occurred by d1, but substantial changes persisted through d7. Metabolites could be classified into those which were predominantly downregulated (e.g., triglycerides, bile acids, cholesterol esters, lysophosphatidylcholines, indoles and derivatives), up- or downregulated (e.g., phosphatidylinositol, phosphatidylethanolamines, phosphatidic acids, ceramides), or upregulated (free fatty acids, monoglycerides). Concentrations of food- and/or microbiota-derived metabolites (indole derivatives, trimethylamine N-oxide, trigonelline) were markedly reduced particularly on d1 and d3. Changes in metabolite concentrations correlated most strongly with plasma C-reactive protein concentration (r = -0.67 to 0.59) and blood leukocyte count (-0.63 to 0.32) and less with intra-operative (-0.62 to 0.5) and ICU care (-0.52 to 0.38) parameters. Of note, neither CRP nor leukocyte count correlated significantly with an intra-operative or ICU parameter.

**Conclusions:** These results reveal pronounced changes in plasma metabolite populations after CABG, which likely result from the combined effects of surgical and post-operative stress, systemic inflammation, reduced dietary intake, and changes in gut microflora.

## INTRODUCTION

Despite advances in interventional therapies and medication, coronary artery bypass grafting (CABG) is still globally considered the gold standard for multivessel coronary artery disease (CAD).^1^ This complex surgical procedure is usually conducted with cardiopulmonary bypass (CPB) using the heart-lung-machine (HLM). The major role of HLM is to support heart and lung function, maintaining blood circulation and oxygenation while heart beat is stopped for the duration of surgery. This is achieved by de-routing the blood through the tubes and membranes of the HLM,^2^ where intimate contact between blood components (cellular and plasmatic fractions) and the non-endothelial surfaces of CPB and a subsequent post-CPB reperfusion injury could lead to aberrant blood cellular responses, i.e. by generating reactive oxygen and nitrogen species.^3^ Such responses are associated with increased post-surgical systemic inflammation with a high likelihood for organ dysfunctions. Furthermore, traumatic incisions made during the surgical procedure are also considered key factors for acute systemic inflammation among these patients.^4^ Though both traumatic surgical injury and CPB have been known to induce conventional pathological factors including complements, endotoxin release, cytokines, oxygen-free radicals, and platelet-activating factors, changes in small-molecule metabolite populations are now being increasingly appreciated as manifestations of distinct inflammatory endotypes.^5^

Metabolic signatures in CAD provide rapid and systematic identification of small-molecule metabolites and distinct lipid metabolites, which are considered as “fingerprint” of an individual, in particular at the time of sampling. Indeed, altered cellular metabolism due to an underlying atherosclerotic pathological condition in CAD has been clearly evidenced by metabolic profiling in blood-based fluids.^6^ Plasma lipidomics includes profiling of sterol lipids, glycerolipids, glycerophospholipids, and sphingolipids, which are considered the most abundant lipids.^7^ Of note, both phosphatidylcholines (PC) and lysophosphatidylcholines (LPC) are important members of glycerophospholipids, where PC are integral molecules for (i) maintaining the structure of cell membrane and (ii) circulating lipoproteins and natural surfactants, and LPC is a hydrolyzed bioactive catabolite of PC. Both molecules play crucial roles in atherosclerosis development and progression.^8^

Bile acids (BAs) are emerging as key metabolites in the context of cardiovascular health and disease, where they are originally synthesized in the liver and further metabolized by gut microbiota.^9^ These BAs play pivotal roles in lipid metabolism, as well as in cardiovascular physiology upon signaling through the bile salt receptor (the farnesoid-X receptor (FXR)).^10^ The BA-FXR signaling axis is known for its anti-atherosclerotic properties, by regulating lipid profiles and vascular tension, and thereby showing protective effects during atherogenesis.^11,12^ Nevertheless, dysregulated BA levels could also contribute to cardiovascular pathologies.^13^ This complexity is further underscored by research linking BAs with gut microbiota in the regulation of myocardial function.^14^ Despite this, the exact mechanisms remain poorly understood, with some studies indicating that altered bile acid profiles, particularly the balance between primary and secondary BAs, may influence cardiac outcomes in heart disease.^15^ Furthermore, clinical data suggest that serum concentration of BAs is inversely correlated with the presence of CAD and myocardial infarction,^16^ highlighting the dual role of BAs in both protection and injury of cardiac tissue.

Likewise, tryptophan (Trp) metabolism plays a significant role in cardiovascular physiology, where kynurenine, one of the Trp catabolites which breaks down further into several biologically active metabolites,^17^ is now considered a biomarker for coronary artery disease.^18, 19^ Furthermore, Trp is also readily catabolized by intestinal microbiota to produce indole and derivatives thereof, including indoleacetic acid, indolealdehyde, indolepropionic acid, and indole sulfate.^20^ Of note, indole derivatives are ligands for the aryl hydrocarbon receptor (AhR), and AhR signaling plays important roles in maintaining vascular homeostasis.^21^ Most of these microbial-generated indole derivatives possess beneficial roles by (i) promoting tolerance; (ii) regulating mucosal integrity and gut permeability; (iii) promoting anticoagulant properties; (iv) ameliorating atherogenesis, showing their cardioprotective effects.^22^ In contrast, indole sulfate accumulates as uremic toxin and therefore has a detrimental role in patients with vascular disease comorbid with chronic kidney failure.^23, 24^ Elevated levels of indole sulfate are involved in the pathogenesis of aortic arterial stiffness among CAD patients.^25^ Though many studies show a causal relationship between blood metabolites and CAD progression, there are still major gaps in knowledge in identifying prominent metabolic signatures during surgery-related acute stress for a progressive time frame. Such metabolites could be regarded as biomarkers to predict postsurgical adverse events and CAD outcome.

We therefore investigated the impact of cardiac surgery and the consequences of using CPB, on plasma metabolite alterations among CAD patients. Here, we analyzed CAD patients before surgery and followed them for three time points post CABG, to observe a possible link between alterations in plasma metabolites and systemic acute inflammation. Furthermore, we focused on associations between metabolic changes and several perioperative parameters including (i) CPB utilized time; (ii) aortic cross clamping time; (iii) mechanical ventilation time; (iv) epinephrine/dobutamine time; and (v) norepinephrine time. The results suggest that these metabolic indicators could serve as valuable molecular tools to identify disturbances in key metabolic pathways among CAD patients who have undergone CABG.

## METHODS

### Study cohort

This prospective study was approved by the Ethics Committee of Otto-von-Guericke-University Hospital (OVGU, Magdeburg, Germany) (file no. 75/19), which is in accordance with the ethical principles of the Declaration of Helsinki. After giving informed consent, N=24 patients with coronary artery disease (CAD) from the Department of Cardiothoracic Surgery at OVGU were enrolled in 2023, all undergoing elective on-pump CABG. Among them, n=3 (12.5%) had two vessel disease, and n=21 (87.5%) had three vessel disease. Inclusion criteria were (i) adults >18 years of age and (ii) elective on-pump CABG. Exclusion criteria were (i) intake of immunosuppressive medication, (ii) viral hepatitis or HIV infection, and (iii) anemia with hemoglobin <7 mmol/l. All patients underwent the same standardized protocol for anesthesia (≥8 hours no food and ≥2 hours no beverage intake before surgery, general anesthesia with propofol, rocuronium and sufentanil), perfusion, and surgery. All patients resumed oral food intake on day 1 post-OP except for two patients who developed a complicated clinical course.

### Study design

Blood samples were collected at the following time points: two days before surgery (d0, n=24), one day post-surgery (d1, n=24), three days post-surgery (d3, n=24), and either six (n=10) or seven days post-surgery (n=10). Due to early recovery, 10 patients were sampled on day 6 post-surgery before discharge to rehabilitation. A two-group comparison between day 6 and 7 post-surgery showed no significant separation in the principal component analysis (PCA) based on metabolic differences, and a differential abundance analysis (fold change ([FC]>|1.5|, false discovery rate [FDR] < 0.05) did not reveal any significantly changed metabolites. Thus, samples from day 6 and 7 post-surgery were collectively analyzed as day 7 (d7, n=20). Metabolite data were not available for 4 patients: one patient died on day 6 due to sudden cardiac death after transfer to normal ward, and the blood sample for metabolomic studies was not taken from 3 patients due to internal error. The data was paired, and each post-surgery sample (d1, d3, d7) was compared to the pre-surgery sample (d0) from the same patient.

### Blood and clinical parameters

Plasma samples were obtained from venous blood and used for metabolite profiling. Blood was centrifuged to sediment cells and debris, and aliquots of supernatants were frozen at -80°C within 2 hours. Additionally, the following laboratory parameters were measured at each time point (d0, d1, d3 and d7) at the central laboratory of OVGU hospital using standard protocols: leukocyte count, C-reactive protein (CRP), alanine aminotransferase (ALAT), creatine kinase (CK), creatinine, quick, international normalized ratio (INR), and activated partial thrombin time (aPTT). Furthermore, the recorded surgical parameters comprised number of vessel bypasses, cardiopulmonary bypass time (CPB time), and aortic cross clamping time (ACC time). The intensive care unit (ICU) parameters that were recorded comprised length of ICU stay, time of total mechanical ventilation (MV time), and length of catecholamine therapy (epinephrine or dobutamine therapy [E/Dob time] and norepinephrine therapy [NE time]).

### Targeted metabolomic profiling

Metabolite concentrations were measured on a SCIEX 5500 QTrapTM mass spectrometer (SCIEX, Darmstadt, Germany) using the MxP Quant 500XL kit (Biocrates, Life Sciences AG, Innsbruck, Austria). The kit combines flow injection analysis tandem mass spectrometry (FIA-MS/MS) for lipids and liquid chromatography tandem mass spectrometry (LC-MS/MS) using Agilent 1290 Infinity II liquid chromatography (Santa Clara, CA, USA) coupled with a tandem mass spectrometer for small molecules. Depending on the sample to be analyzed, this kit allows the quantification of up to 1019 metabolites: alkaloids (1), amine oxides (1), amino acids (20), amino acid related molecules (30), bile acids (14), biogenic amines (9), carboxylic acids (7), cresols (1), fatty acids (12), hormones and related metabolites (4), indoles and derivatives (4), nucleobases and related metabolites (2), vitamins and cofactors (1), carbohydrates and related metabolites (1), acylcarnitines (40), lysophosphatidic acids (8), phosphatidic acids (41), lysophosphatidylcholines (12), phosphatidylcholines (76), lysophosphatidylethanolamines (43), phosphatidylethanolamines (95), lysophophatidylglycerols (10), phosphatidylglycerols (64), lysophosphatidylinositols (16), phosphatidylinositols (53), lysophosphatidylserines (12), phosphatidylserines (18), sphinganines and sphingosines (8), sphinganine and sphingosine phosphates (8), sphingomyelines (15), ceramides (29), dihydroceramides (8), glycosylceramides (34), cholesterol esters (22), monoglycerides (12), diglycerides (44), and triglycerides (242). Metabolite extraction and all analytical assays were conducted in accordance with the protocols provided by the manufacturer (https://biocrates.com/mxp-quant-500-xl/, accessed on 02 Mai 2024). The WebIDQ ™ software tool (Biocrates Life Science AG, Innsbruck, Austria) was used for peak integration and to calculate metabolite concentrations.

### Quality screen

The samples were measured on three 96 well plates. To assess metabolic changes as accurately as possible, all metabolites were included in the analyses except for those with a constant value across all samples in the respective comparison. 51, 49 and 52 metabolites were excluded when comparing d1 vs. d0, d3 vs. d0 and d7 vs. d0 respectively. **Table S1** shows a complete list of included metabolites in each comparison. Any concentrations that were measured below limit of detection (LOD) were replaced with the pseudovalue LOD/2.

### Statistical analyses

Due to non-normal distribution of the data, nonparametric statistical tests were employed. For principal component analysis (PCA), data were log10 transformed to correct for heteroscedasticity and auto-scaled to make all variables equally important.^26^ Statistical Analysis of Metabolomics Data. University of North Carolina at Charlotte; Department of Bioinformatics & Genomics. Retrieved April 21, 2024, from https://www.uab.edu/proteomics/metabolomics/workshop/2014/statistical%20analysis.pdf). Mean Euclidian distances were calculated for each metabolite class based on the first two principal components of the PCAs in the respective comparison to assess the metabolite classes with the greatest degree of change. Differential abundance analysis was performed using the Wilcoxon rank sum test to assess significance of differences between groups based on paired samples. The Benjamini-Hochberg correction was implemented with a false discovery rate (FDR) of 0.05 to account for multiple testing. For paired analysis, fold change (FC) was determined by calculating the ratio between paired samples, resulting in one FC per pair. The means of these FC values, pair means, was then computed. A FC threshold of ≥|1.5| was set for differential abundance analysis. Spearman’s rank correlation coefficient was used to assess correlations. PCA, differential abundance analysis, and correlation testing were performed using the open-source software MetaboAnalyst 5.0 (https://www.metaboanalyst.ca/). Values for standard parameters that were <LOD were replaced by LOD/2: i.e., CRP LOD = 0.6 mg/l, was replaced with 0.3 mg/L.

## RESULTS

### Patient cohort

Demographic and clinical characteristics of the patient cohort are summarized in **Tables 1** and **2**. There was the expected preponderance of male patients and high proportion of patients with comorbidities which confer an increased cardiac risk (diabetes, overweight, COPD) (**Table 1**). Most patients presented for bypass surgery for 3 or more coronary vessels, and the recorded values for bypass time, need for vasopressors, and length of ICU stay fell into the expected range. Postoperatively, there was a transient increase in creatine kinase (indicating cardiac ischemia), but renal and liver function parameters remained stable (**Table 2**). There were no significant sex-specific differences except a higher blood leukocyte count in women on post-op day 3 (12.1 (10.1-14.8) vs. 9.6 (7.13-13.5), *p* = 0.024). Median CRP values rose greatly, manifesting the typical peak on day 3 and subsequent decline, but the median value of 59.5 mg/dL on d7 indicated ongoing systemic inflammation. In contrast, the marked leukocytosis on d1 essentially normalized by d7. Taken together, these data suggest that the patient cohort has the typical clinical characteristics of individuals presenting for elective coronary bypass graft surgery and manifests the expected postoperative evolution of inflammatory parameters.

**Table 1.** Baseline and intra-operative characteristics.

| **Table 1. Baseline and intra-operative characteristics** |  |
| --- | --- |
| **Demographic characteristics** |  |
| Gender female, n (%) | 8 (33.3) |
| Age at first blood collection, years (median [range]) | 66 (49-82) |
| BMI (median [range]) | 28.2 (23.1-39.7) |
| Acute coronary syndrome, n (%) | 4 (16.7) |
| LVEF, % (median [range]) | 55 (18-65) |
| **Comorbidities** |  |
| Diabetes mellitus (I or II), n (%) | 12 (50) |
| Smoker, n (%) | 8 (33.3) |
| COPD, n (%) | 2 (8.0) |
| Chronic kidney disease (eGFR <90 ml/min/1.73m<sup>2</sup>), n (%) | 18(75) |
| Atrial fibrillation, n (%) | 4 (16.7) |
| **Surgical procedure** |  |
| Double Bypass, n (%) | 2 (8.2) |
| Triple Bypass, n (%) | 11 (45.8) |
| > Triple Bypass, n (%) | 11 (45.8) |
| **Surgical parameters** |  |
| Cardiopulmonary bypass time, min (median [range]) | 83.5 (46-166) |
| Aortic cross clamping time, min (median [range]) | 51 (32-81) |
| Operating time, min (median [range]) | 200 (105-315) |

**Table 2.** Hematologic and clinical chemistry parameters.

| **Table 2. Hematologic and clinical chemistry parameters** |  |  |  |  |  |
| --- | --- | --- | --- | --- | --- |
| **Parameter** | **d0** | **d1** | **d3** | **d7** | **Reference values** |
|  | **Median (range)** |  |  |  |  |
| Leukocyte count (Gpt/L) | 7.7 (5.8-93.8) | 13.4 (5.2-17.7) | 10.7 (7.1-19) | 9 (3.2-20) | 3.9 - 10.4 |
| CRP (mg/L) | 1.4 (0.3-67.1) | 32.7 (13.8-114.0) | 131.3 (58.6-242.5) | 59.5 (22.7-169.1) | < 5 |
| ALAT (μmol/s.L) | 0.5 (0.3-1.0) | 0.4 (0.3-2.5) | 0.5 (0.2-1.5) | 0.6 (0.0-1.6) | 0.17 - 0.58 |
| Creatin kinase (μmol/L) | 2.2 (0.5-10.5) | 8.2 (3.4-74.2) | 4.8 (0.8-35.4) | 2.1 (0.6-18.0) | < 0.40 |
| Creatinine (μmol/L) | 80.5 (60.0-232.0) | 73.5 (0.4-227.0) | 78 (57.0-252.0) | 79.5 (59.0-234.0) | 45 - 84 |
| CKD-EPI (ml/min/1.73m<sup>2</sup>) | 1.4 (0.4-9.3) | 1.5 (0.4-23.3) | 1.4 (0.2-1.8) | 1.3 (0.2-1.9) | ≥ 1.5 |
| Quick (%) | 102.0 (76.0-120.0) | 84.0 (32.3-110.0) | 87.0 (73.0-104.0) | 83.5 (74.0-96.0) | > 70 |
| INR | 1.0 (0.4-10.0) | 1.1 (1.0-1.4) | 1.1 (1.0-1.2) | 1.1 (1.0-1.2) | < 1.15 |
| aPTT (sec) | 28.7 (25.7-34.2) | 27.8 (19.4-32.9) | 28.8 (24.6-39.7) | 29.4 (23.3-58.0) | < 34.4 |

### Postoperative changes in metabolite populations

A PCA was performed to assess global differences in metabolite populations at each of the 3 post-operative time points (d1, d3, d7) with respect to the preoperative samples (d0). While substantial separation was already evident on d1, it increased further by d3 and persisted through d7. However, variance shifted from PC1 to PC2 on d7, indicating that it was driven, in part, by different metabolites than at the earlier time points. At all three time points, the greatest degree of variance with respect to d0 was due to alterations in triglyceride (TG) concentrations. For instance, the 20 analytes with the greatest contribution to variance in the first component (PC1, which contributed 17-23.5 % of overall variance) were TGs at all-time points, indicating a considerable degree of coregulation within this analyte class (**Figure 1A-C, Table S2)**. Regarding PC2, PA and PG contributed most to variance at all-time points, peaking on d3 and decreasing on d7. TG impacted on PC2 only on d1. MG and LPEs contributed to variance in PC2 only on d7, largely explaining the separation between d0 and d7 samples along PC2 at this time point.

**Figure 1.**
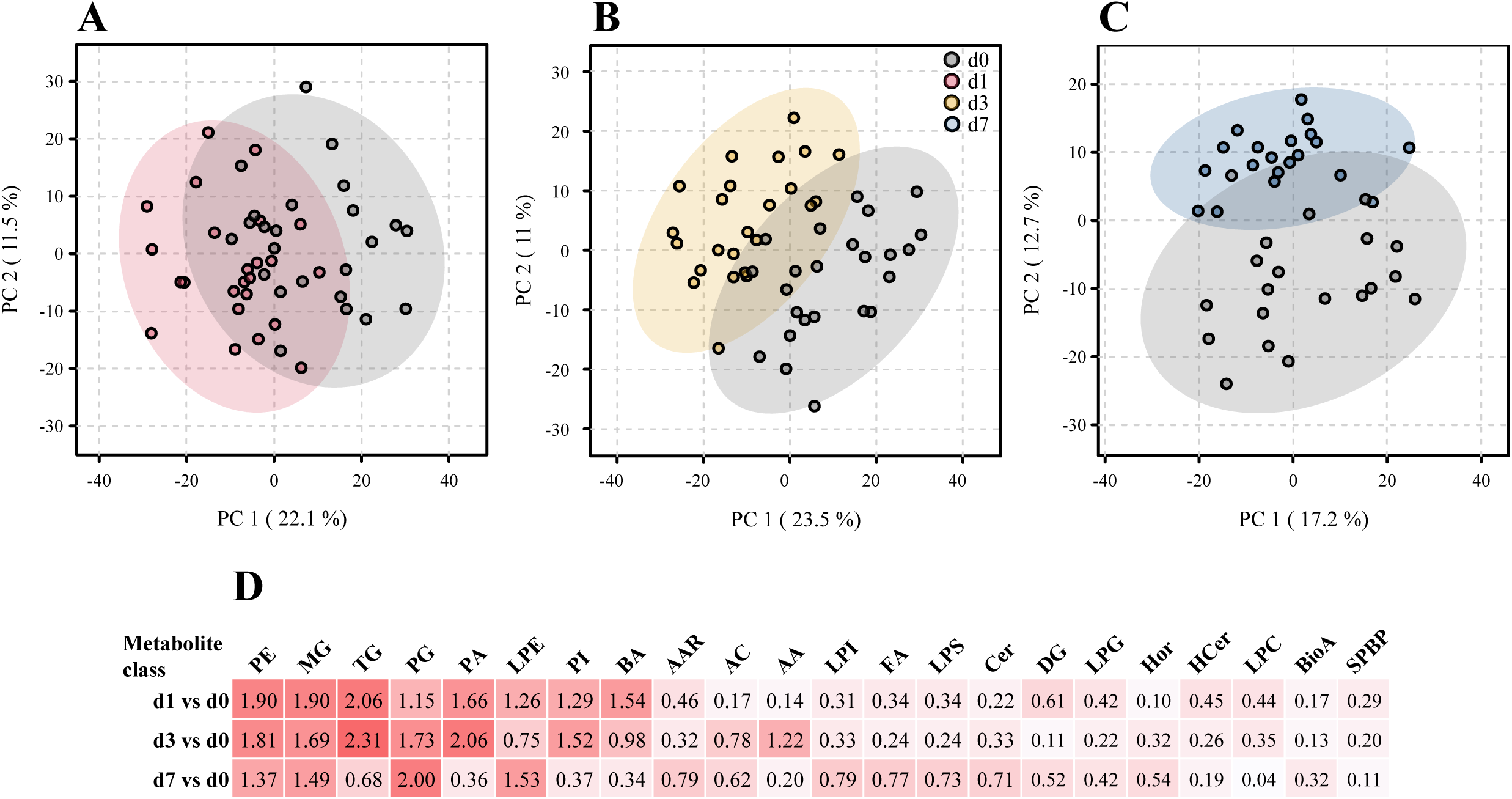
Global and class-specific changes in metabolite populations at the three time points post-op. A principal component analysis (PCA) was performed using 968, 970 and 967 metabolites that passed quality assessment for the comparisons d1 vs. d0, d3 vs. d0 and d7 vs. d0, respectively (Table S1). The degree of reprogramming within individual metabolite classes was additionally assessed by Euclidian distance analysis. Total patient numbers (N) = 24 with four time points/patient, where N = 24 (d0); 24 (d1); 24 (d3); 20 (d7). **A-C,** PCA; **D,** Euclidian distance analysis. A complete list of Euclidian distances across all metabolite classes and the abbreviations used is contained in **Table S3**.

An Euclidian distance analysis supported the above notion that the greatest changes in metabolite populations occurred by d1 and d3, whereby the greatest changes were seen in lysoPEs, PEs, TGs, MGs, PC, PAs, lysoPC, and bile acids. The latter stood out in that there was a marked increase in variance on d1, which decreased by d3 and essentially normalized by d7. Nonetheless, several metabolite classes showed the largest Euclidian distance on d7. These were characterized by, overall, moderate Euclidian distances on d1 and d3 (e.g., LPI, FA, LPS, Cer), underscoring the differences in metabolite populations on day 7 compared to d1 and d3.

A hierarchical clustering analysis based on the 100 most differentially abundant metabolites revealed a high degree of separation between the pre- and post-operative samples at all three time points, whereby all postoperative samples clustered in their own clade only on d1 (**Figure 2**, **Figures S1**). There was an overall tendency towards lower analyte concentrations on d1, but MGs constituted a notable exception in that concentrations of the three MG among these 100 analytes were higher post-op. Furthermore, this analysis revealed a strong coregulation of metabolites within one class, es exemplified by the clades 1-4, marked by a red circle, containing mostly BA (clade 1), TG (clade 2), PC/LPC (clade 3), and PE (clade 4). Co- regulation within metabolite classes (particularly within PS, PI, PG, PE, PA, TG, LPC, CE, BA, and AA) is also evident in a matrix based on correlations among all metabolites (**Figure S2)**. This analysis also revealed between-class correlations, for instance of PI with PG and PA and of TG with PC, a subgroup of LPC, and DG.

**Figure 2.**
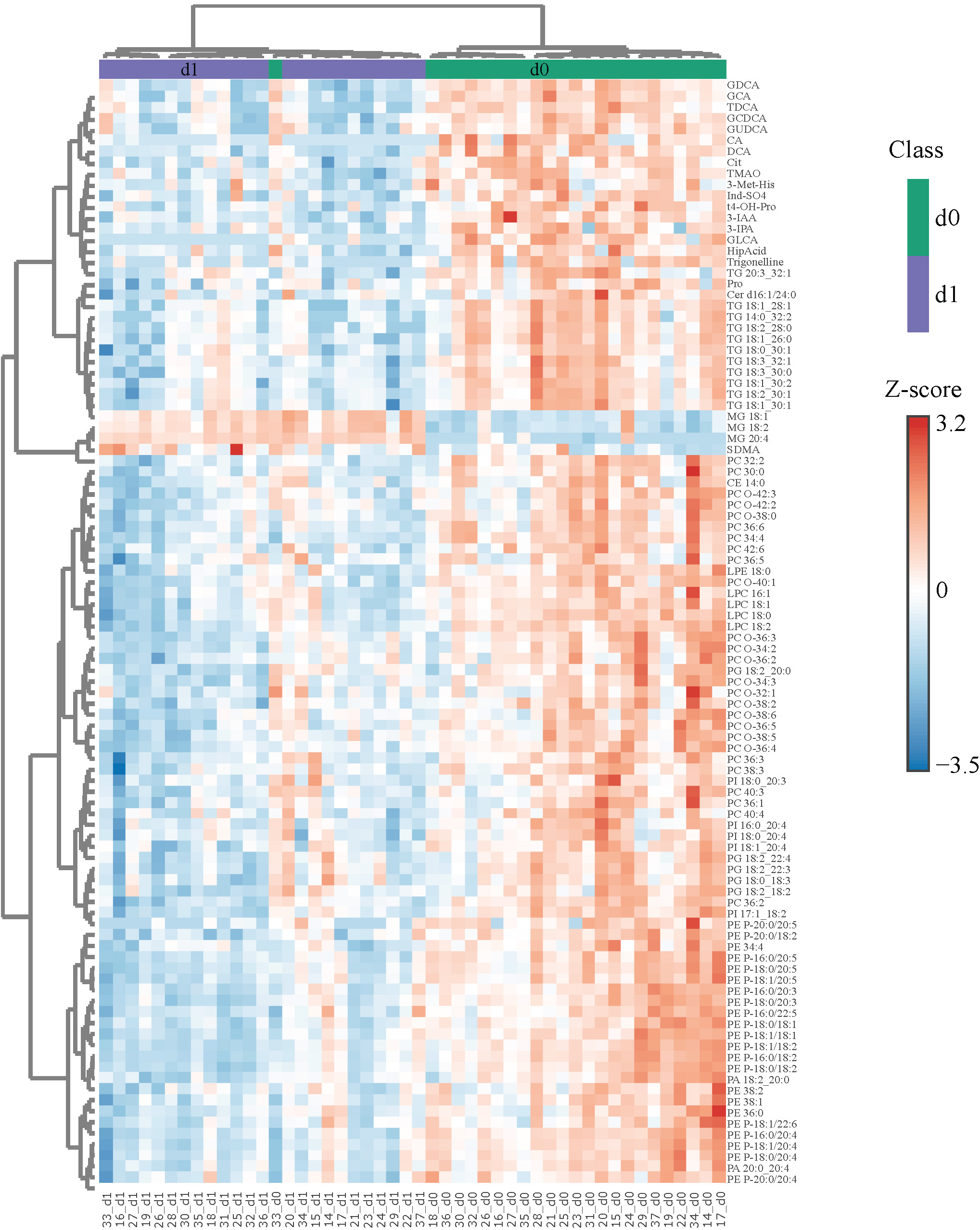
Differences in metabolite concentrations distinguish nearly perfectly between d0 and d1 samples. The 100 most significant (FDR obtained with t-test = 1.3e^-9^ – 5.86e^-5^) differentially abundant metabolites were selected by Euclidean distance for d1 with respect to d0 and subjected to unsupervised hierarchical biclustering analysis. Each colored cell in the heatmap corresponds to the group average concentration of the analyte with respect to the mean-centered and divided by standard deviation of the analyte (z-score). Y-axis = metabolite dendrogram and N = 24 (d0) and 24 (d1).

### Differential abundance analysis

Results of a differential abundance (FC ≥|1.5|, FDR <0.05) analysis are visualized in **Figure 3** and summarized in **Table 3**. As seen in the volcano plots (**Figure 3A-C**) and **Figure S3** there was a trend towards higher *p* values and FC (log10) values from d1 to d7. The highest number of differentially abundant metabolites was observed on d3, but even on d7 there were still 154 differentially abundant metabolites. Overall, most of the differentially abundant metabolites were downregulated. However, the number of downregulated metabolites decreased throughout the time course, whereas the number of upregulated metabolites increased, which resulted in a slight preponderance of upregulated vs. downregulated metabolites by d7 (**Table 3**).

**Figure 3.**
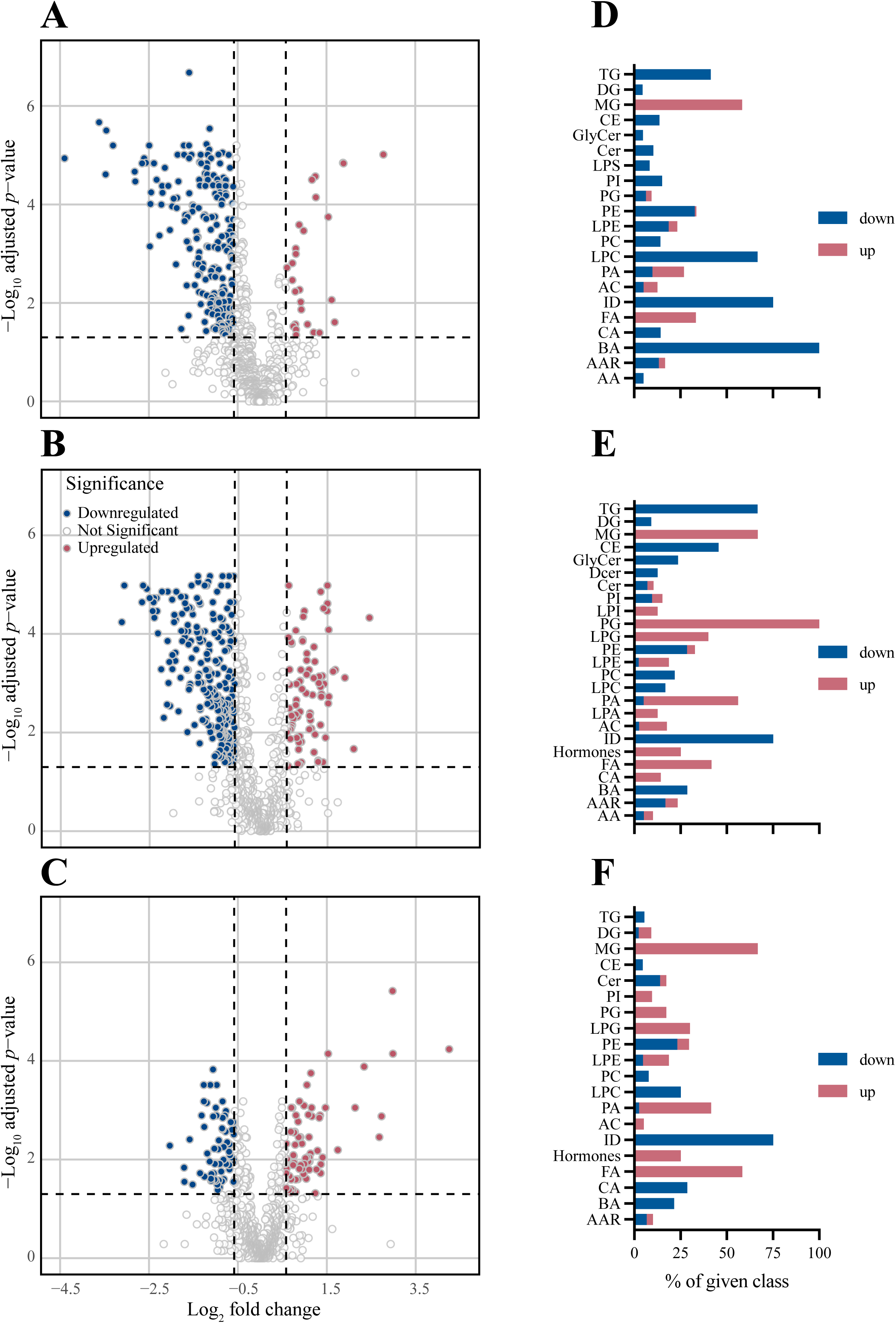
Differential abundance analysis based on the same metabolites as figure 1 reveals a shift from down- to upregulation during post-operative recovery. Differentially abundant (FC ≥|1.5|, FDR <0.05 obtained with Wilcoxon rank-sum test) metabolites with respect to d0 are indicated for each time point post-op. For each day, a volcano plot indicating differential abundance of individual metabolites and a bar chart indicating the direction of abundance change in each metabolite class are shown. Blue bars = downregulation; red bars = upregulation. **A,D,** d1. **B,E**, d3. **C,F**, d7 and N = 24 (d0); 24 (d1); 24 (d3); 20 (d7).

**Table 3.** Number (%) of differentially abundant metabolites post-op.

| **Table 3.** Number (%) of differentially abundant metabolites post-op |  |  |  |
| --- | --- | --- | --- |
| **Day post-op** | **Differentially abundant metabolites** |  |  |
|  | **Total no.** | **No. (%) up** | **No. (%) down** |
| d1 | 262 | 34 (13%) | 228 (87%) |
| d3 | 334 | 82 (25%) | 252 (75%) |
| d7 | 154 | 82 (53%) | 72 (47%) |

Metabolite classes could be grouped into two kinetic patterns: those that were consistently downregulated (TG, BA, CE, ID) and those that were consistently upregulated (FA and MG) (**Figure 3D-F**, **Figure 4**, **Figure S4 and 5**). Notably, all BA were downregulated on d1 and 35, 7 % remained downregulated more than 1.5-fold on d3 and d7. Likewise, LPC and ID were consistently downregulated. Downregulation of TG was most pronounced on d3 (66%), but nearly normalized by d7. Substantial persistent upregulation was observed with FA and MG. Of the metabolite groups with few or only a single member, substantial downregulation of three metabolites was noted. (i) Concentrations of the alkaloid trigenolline were very low on d1 and reached only about 25% of pre-op levels by d7. Trigenolline is found in foods, notably in coffee beans, where it makes up about 1% and can also originate from gut microflora ^27^. (ii) Similar downregulation was observed with trimethylamine N-oxide (TMAO), which can originate from gut microbiota or hepatic metabolism of trimethylamine. TMAO is closely related to the so called “western diet”, which is rich in choline and carnitine, and is associated with cardiovascular disease.^28^ (iii) 3-IAA is a Trp metabolite which is a common growth hormone in plants, but also an intermediate of Trp catabolism in humans. Thus, the initially low, but then rising plasma levels of trigenolline and TMAO likely reflect absence of oral alimentation on d1, and the gradual resumption of an oral diet thereafter. There was a mild-to-moderate increase in cortisol post-op (indicating a physiologic stress response), whereas cortisone levels did not increase.

**Figure 4.**
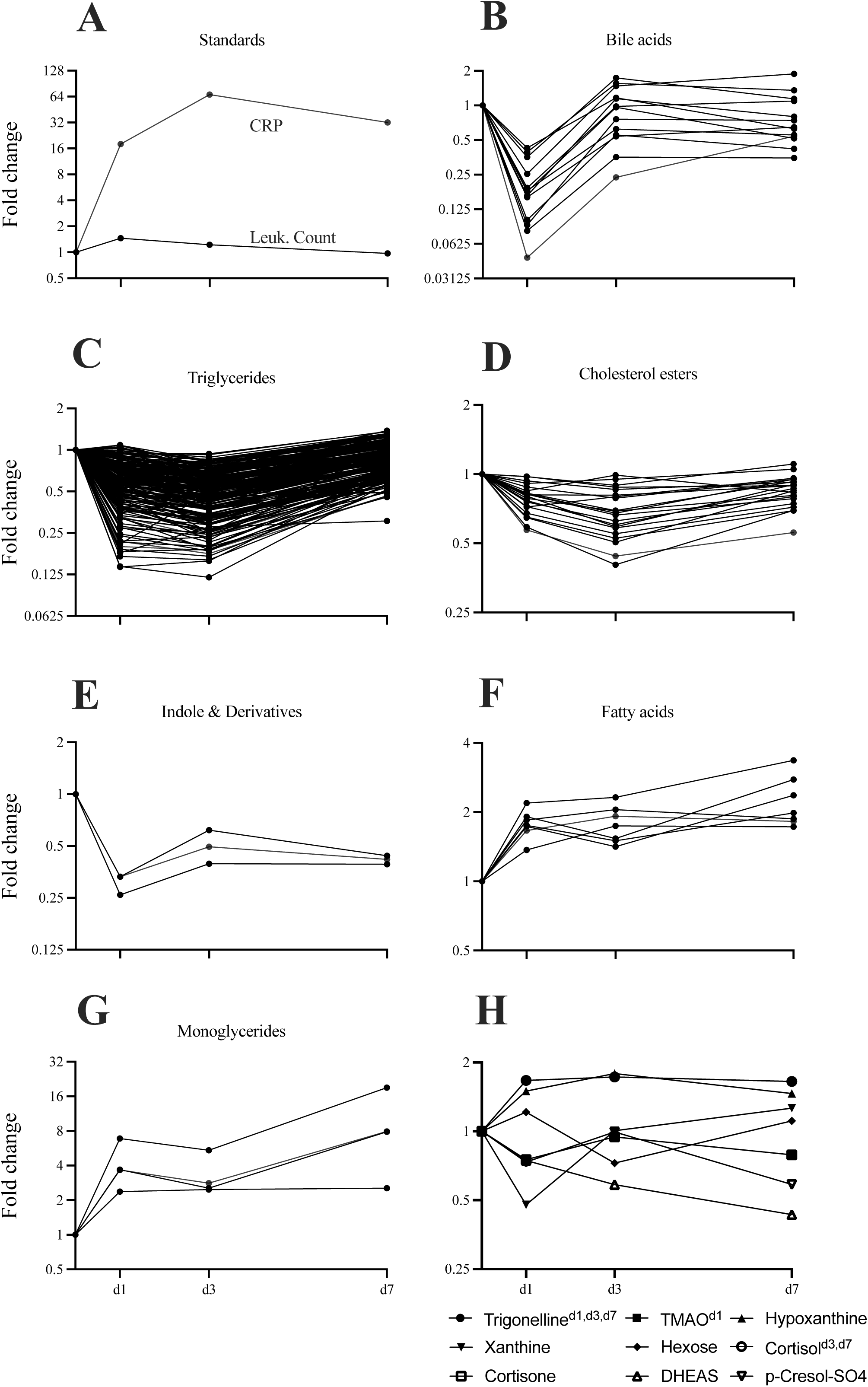
Kinetics of concentration changes in selected metabolite classes and blood inflammatory parameters throughout the time course. Concentration dynamics are expressed as fold change with respect to d0. **A,** Blood CRP and leukocyte count. **B-H,** Plasma metabolites. **B-E**, Selected predominantly downregulated metabolite classes (bile acids *n* = 13, triglycerides *n* = 238, cholesterol esters *n* = 22, indoles and derivatives *n* = 3). **F,G,** Selected predominantly upregulated metabolite classes (fatty acids *n* = 8, monoglycerides n = 11). **H**, Selected individual metabolites of interest (trigonelline, TMAO, cortisol, cortisone, DHEAS). Days on which fold change was significant with respect to d0 are indicated as superscripts. N = 24 (d0); 24 (d1); 24 (d3); 20 (d7).

### Correlations between clinical parameters and metabolites

We then aimed to identify clinical parameters that are most closely associated with changes in metabolite abundance and can be considered the main drivers of the marked metabolic alterations post-op. Among the intra-operative variables, the most significant correlations with metabolites at all three time points were observed for NE time, CPB time, and E/Dob time (**Figure 5A, Figure S6, Table 4**). However, most significant correlation coefficients were of only modest strength, ranging between 0.25-0.5. On the other hand, substantially stronger correlations were observed between the markers of inflammation (CRP and leucocyte count) and metabolites, indicating that systemic inflammation is a strong, but certainly not the only, driver of the observed metabolite alterations. We then tested whether we could identify intra-operative parameters responsible for the observed systemic inflammation. However, the only significant correlation with inflammation was between ALAT and leukocytes (*r*= -0.41, *p*= 0.0006) (**Figure 5B**).

**Figure 5.**
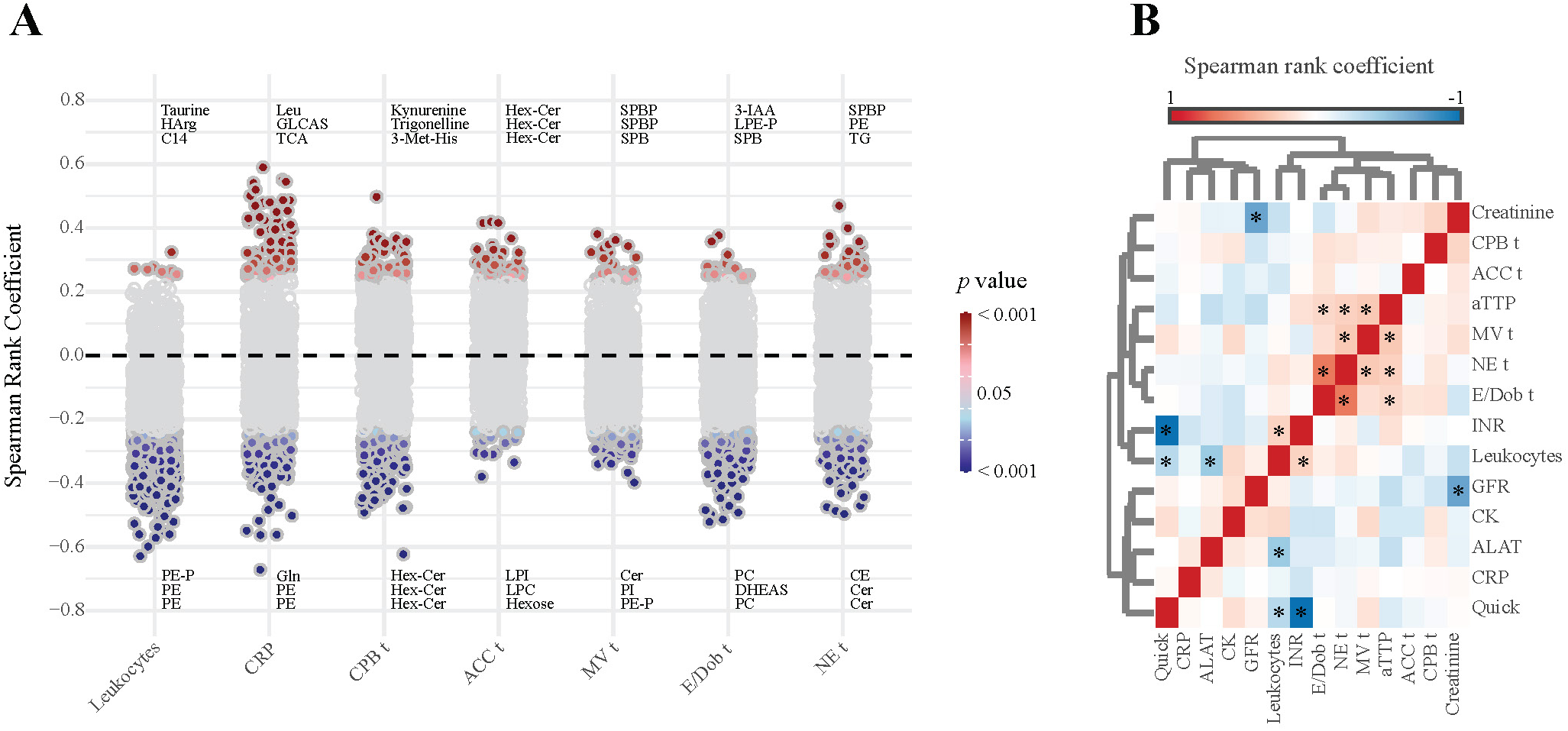
Correlations between intra-operative parameters, inflammation markers, and metabolites, based on Spearman rank coefficients. **A**, Correlation with metabolites, pooled analysis of data from all three time points combined. Each circle indicates one metabolite. Given metabolites represent the ones with highest correlation coefficient. Grey fill color = *p* > 0.05. **B,** Correlations among intra-operative parameters (n = 5), inflammation markers (n = 2), and clinical chemistry parameters (n = 6). * = *p* < 0.05. *P* values based on FDR obtained with t-test. N = 24 (d0); 24 (d1); 24 (d3); 20 (d7).

**Table 4.** Number of significant correlations (*p* ≤0.05) between metabolites and clinical parameters.

| Parameter <sup>a</sup> | d1 | d3 | d7 | Sum |
| --- | --- | --- | --- | --- |
| CRP | 152 | 32 | 54 | 238 |
| Leukocyte count | 66 | 123 | 29 | 218 |
| E/Dob time | 72 | 63 | 46 | 181 |
| NE time | 85 | 37 | 46 | 168 |
| CPB time | 56 | 64 | 47 | 167 |
| MV time | 53 | 21 | 27 | 101 |
| ACC time | 34 | 20 | 27 | 81 |
| All | 518 | 360 | 276 | 1154 |

<sup>a</sup> Arranged in descending order according to sum of significant correlations
Abbreviations: CRP, C-reactive protein; E, Epinephrine; Dob, Dobutamine; NE, Norepinephrine; CPB, Cardiopulmonary bypass; MV, Mechanical ventilation; ACC, Aortic cross clamping

## DISCUSSION

Our study is the first to show significant plasma metabolite changes after CABG in both acute and recovery phase, distinguishing it from similar studies.^29, 30^ Although changes in metabolites were evident during the acute phase and normalized during the recovery phase, intriguingly, there were few metabolite classes, belonging to the lipid, bile acid, and Trp derivatives, which exhibited a steady kinetic pattern throughout our investigations. Notably, plasma TG levels were significantly downregulated and were associated with increases in both MG and FA, suggesting perioperative modulation in lipid metabolism. Such alterations in metabolites could occur due to several plausible factors, including surgical trauma-associated inflammatory milieu, fasting, catecholamine-induced lipolysis, and heparin treatment. Of note, disturbances in gut microbiota community also play a significant role in inducing changes in metabolite profiles.^31^

Increased systemic inflammation is a well-known consequence of CABG, in the process of which macrophages with inflammatory^high^ phenotype tend to accumulate plasma-circulating TG, by either endocytosis or receptor-mediated internalization.^32^ Such TG-laden inflammatory macrophages exhibit reduced capacity for lipolysis, which further lowers the inflammatory attributes of macrophages associated with increased mitochondrial dysfunction, decreased ability to polarize, migrate,^33^ and phagocytose, and reduced PGE2 production.^34^ This suggests a negative feedback loop to minimize the inflammatory state of these macrophages upon intracellular TG accumulation and could explain the reduced post- operative plasma TG levels. On the other hand, TG, CE, and PC accumulate in intracellular lipid droplets as cells respond to stress.^35–37^ In our cohort, surgery-induced acute stress could promote lipid droplet turnover to compensate the energy and redox imbalances, where TG are constantly being hydrolyzed (via lipolysis or lipophagy) to convert into free FA and MG,^38^ as reflected by their consistent upregulation in postoperative CAD circulation. Of note, CE-mediated lipid droplet nucleation is facilitated by these intracellular accumulated TG, since reduced plasma CE were also evident in our cohort.^39^ Another contributing factor is the state of fasting, where the entire patient cohort remained fasting ≥6 hours prior to surgery until an oral diet was resumed on the first postoperative day. In addition to cardiac surgery stress,^40^ this nutrient stress also triggers the sympathetic nervous system^41^ to secrete catecholamines (norepinephrine) that activate lipolysis to satisfy the cellular energy demand, by promoting the breakdown of stored TG into FA and MG.

Furthermore, every patient received high-dose intravenous (IV) unfractionated heparin (400 IU/kg body weight) before commencement of extracorporeal circulation, followed by low-dose prophylactic therapy until discharge. Since IV administered heparin rapidly increases lipid hydrolyzing enzymes (lipoprotein lipase), there could be a massive hydrolysis of TG into FA and MG.^42^ Likewise, the entire patient cohort were uniformly treated with opioids, as peri- and post-operative pain management medications. Opioid could be regarded as a significant driving factor for changing cellular metabolism.^43^ In line with this, the opioid doses could tend to increase cellular lipolytic activity, which could also, in part, generate FA^44^ and MG from intracellular TG deposits to counterbalance increased cellular energy expenditure.^45^

PC, as predominant class of plasma lipids, were consistently decreased in circulation, possibly due to their role in lipid droplet biogenesis^46^ for high-energy supply during surgery-induced inflammation. Another explanation could be PC uptake via the CD36 scavenger receptor, increasing the cellular PC pool in differentiating macrophages, which may accelerate PC turnover and amplify inflammation.^47^ Additionally, monocytes, as macrophage precursors, feature a higher intracellular PC content, particularly with aging.^48^ These concepts hold true in our cohort settings that are representative of an aged population together with ongoing post-surgical inflammation. We therefore propose that an increased demand for intracellular PC content might reflect decreased circulating PC in the blood stream. A study of plasma lipid changes in community-acquired pneumonia (CAP) reported lower PC concentrations during acute presentation, which normalized with clinical recovery.^49^ These results agree well with ours, as acute CAP features pronounced systemic inflammation of a similar magnitude as our cohort. Interestingly, a reduced plasma PC/PE ratio has led to increased levels of NLRP3, an integral component of the inflammasome activation pathway, which promoted changes in left ventricle geometry among CAD patients with insulin resistance.^50^ Since 50 % of our CAD patients have diabetes as comorbidity, a notable decrease in plasma PC/PE ratio could possibly pose an increased risk to these patients in terms of cardiac remodeling. Similar to our findings, plasma levels of PC were also reduced in patients with other chronic inflammatory diseases, such as Alzheimeŕs disease^51^ and diabetic nephropathy,^52^ where both plasma PC and PC-Os were inversely associated with renal-related clinical covariates (eGFR and Albuminuria), ESRD and all-cause mortality in diabetic nephropathy.^52^ Sometimes, choline-deficient diet could also explain the reasons behind a sudden drop in plasma PC levels^53, 54^ among CAD patients. However, a significant decline in plasma lipids could explain an ongoing catabolism due to acute stress,^55^ induced by surgical trauma. Further, these circulating PC might have sequestered by binding to structurally-altered CRP^56^ due to massive inflammation. Since PC is further cleaved by phospholipase A2 activity to generate LPC,^8^ we noticed substantially reduced plasma LPC in CAD patients, which could reflect a limited availability of its precursor molecule, PC, in the circulation. Similar to our findings, plasma LPC levels were decreased among patients with obesity and diabetes^57^ as well as Alzheimeŕs patients with chronic inflammation.^58^ Since LPC act as a carrier molecule for PUFAs,^58^ a deficit of plasma LPC pool might limit supply of PUFA to vascular health, causing vascular dysfunction,^59^ increased inflammation and all-cause mortality.^60^

Several reports have shown alterations in gut microbiota composition following cardiac surgery together with CPB usage.^61, 62^ Such gut dysbiosis led to decreased LPC generation which was, in particular, associated with reduced colonization of *Bacteriodes* population, as investigated in chronic pathologies such as Alzheimeŕs disease^63^ Furthermore, we speculate that there was increased ferroptosis with decreased plasma LPC^63^ in our patient cohort, as iron accumulation and lipid peroxidation have been reported following cardiac surgical procedures, including the usage of CPB.^64, 65^ In line with the above-mentioned beneficial role of PC and LPC, our study shows a negative correlation between PC and PC-Os as well as LPC and acute inflammatory markers (CRP and leucocyte count) and the use of CPB during surgery, highlighting a possible role of PC and LPC in regulating ongoing inflammation.

Changes in plasma bile acids were also noticed in our CAD cohort, where BA metabolites were consistently downregulated, especially on postoperative day 1 and 3. This could reflect limited generation of BA metabolites due to gut dysbiosis,^66^ which was also evident in patients with other chronic inflammation like ulcerative colitis.^66^ Hence, to this extent, there is a high likelihood for a surgical trauma-induced gut dysbiosis in decreasing circulating BA metabolites postoperatively CAD. In fact, increased BA concentrations in the circulation have been reported to modulate blood monocyte function upon specific activation of bile acid receptor (TGR5). These monocytes exhibit decreased phagocytic capacity and reduced production of pro-inflammatory cytokines^67^ during bacterial infection. Nevertheless, bile receptor TGR5 agonism induces nitric oxide (NO) production and hampers the adhesion of circulating monocyte to vascular endothelium.^67^ In line with these reports, we supposed whether bile acid synthesis was reduced or their bioavailability was diminished due to their consumption at cell-expressed receptor binding sites in our cohort. But, in our previous study,^68^ we demonstrated IL-6^high^ producing blood mononuclear cells among postoperative CAD patients, explaining the existence of functional monocytes and therefore, the possibility for decreased BA synthesis. Thus, a deficit in bile acid production could pose an increased risk for amplified inflammation in our postoperative cohort. Intriguingly, BA were positively correlated with CRP in these patients, suggesting a counteracting role of bile acids to reduce ongoing inflammation. This notion was supported by a report showing that increased CRP levels in LPS-induced endotoxemia among healthy volunteers correlate with a rise in total BA concentration^69^ where these increased circulating BA metabolites exhibited immunosuppressive properties. However, a functional relationship between CRP and circulating BA would be interesting to investigate in our CAD cohort. Interestingly, a study on the effect of fasting and refeeding on plasma BA metabolites in mice demonstrated that both primary and secondary BA were decreased during fasting and this condition was reversed upon refeeding. These alterations were largely associated with decreased abundance of BA-metabolizing *Lactobacillus* and *Bifidobacterium* in gut. In addition, the genes involved in hepatic BA synthesis (CYP7A1, CYP7B1, and AKR1D1) were repressed during the fasting state. However, BA metabolizing microbiota as well as hepatic BA synthesizing genes were expressed during re-feeding.^70^ A similar mechanism might underlie the reduced plasma BA levels in our CAD patients, who were also in the state of fasting and gradual refeeding during the peri-and post-operative period.

Next, a consistent downregulated pattern of indole derivatives (indole, 3-IAA, 3-IPA and Ind-SO_4_) was noticed. These molecules are generated as Trp catabolites by the gut microbial community,^22^ are absorbed by intestinal epithelium and enter the blood stream. They are particularly known to fortify intestinal epithelial barrier function, by increasing the expression of tight junctional proteins, and thereby to prevent leaky gut.^22^ Post-surgical gut microbial dysbiosis could explain the reason for reduced plasma indole derivatives in our cohort. To support our findings, significantly reduced levels of indole derivatives, in particular 3-IAA and 3-IPA, have been reported in patients with severe atherosclerosis,^71^ indicating their anti-inflammatory role.^72^ Additionally, 3-IPA is known to enhance systemic homeostasis with the gut-multiorgan axis during acute systemic inflammation.^73^ Although several indole derivatives promote inflammation-attenuating host responses, indole sulfate is considered to be the notable exception, which is a uremic toxin and accumulates in serum of chronic kidney disease patients.^74^ In line with this and despite reduced levels of indole sulfate in our cohort, a positive correlation between indole sulfate and CBP time was noted, reflecting its possible role in oxidative stress. In addition to this, another Trp catabolite (kynurenine) and trigonelline were positively associated with CBP time. Due to their immune suppressive qualities, both kynurenine and trigonelline could be involved in regulating the stressful inflammatory milieu during cardiac surgery. However, specific functions of these metabolites should be investigated.

Taken together, despite the limited number of CAD patients, our current study has demonstrated discrete changes in plasma metabolite populations during the postoperative period. Although we did not observe any distinct pattern of metabolite alterations observed with CBP usage, we presume that surgery-related acute inflammation, in general, plays a significant role in aberrant regulation of metabolites. According to the observation, an excessive generation of free fatty acids in our cohort could pose an increased severity to coronary atherosclerosis, especially among those who are comorbid with T2DM and obesity,^75^ by inducing endothelial dysfunctions and defects in insulin signaling^76^ and sensitivity.^77^ Further PC supplementation as prophylactic measures might be an ideal option for CAD patients prior to cardiac surgery in order to compensate for reduced postoperative PC and LPC levels. A targeted investigation is needed to obtain data on gut microbial communities in CAD patients before and after surgery, especially during fasting and refeeding. A better understanding of depleted microbial taxa is essential to enrich the surgery-scheduled CAD patients with probiotics containing the target microbiota. Such microbial recolonization might benefit these patients to regulate ongoing acute inflammation by maintaining the levels of BA and indole derivatives, which were hampered in postoperative CAD patients. Moreover, a subsequent validation study in similar CAD settings would strengthen the validity of our findings.

## Data Availability

Yes, the data will be available upon request

## AUTHOR CONTRIBUTION STATEMENTS

FN was involved in concept development, experimentation, data analysis and manuscript drafting; MW was involved in concept development, blood sampling and clinical data handling, manuscript drafting; SS was involved in experimentation and manuscript proof-reading; SV was involved in blood sampling and manuscript proof-reading; GA was involved in blood sampling and manuscript proof-reading; JW was involved in manuscript proof-reading; FP provided funds and resources and was involved in concept development, data analysis, and manuscript drafting and finalizing; PV was involved in concept development, experimentation, manuscript drafting and finalizing.

## ACKNOWLEDGEMENTS

We extend our thanks to Ms. Elena Denks for her excellent technical assistance, Esther Meyer for collecting patient blood and organizing the screening for patients, and Dr. Fakhar Waqas (TWINCORE) for support.

